# Structured Codes and Free-Text Notes: Measuring Information Complementarity in Electronic Health Records

**DOI:** 10.1101/2024.10.28.24316294

**Authors:** Tom M Seinen, Jan A Kors, Erik M van Mulligen, Peter R Rijnbeek

**Affiliations:** Department of Medical Informatics, Erasmus University Medical Center, Rotterdam, The Netherlands

**Keywords:** Natural Language Processing, Named Entity Recognition, Clinical Concept Extraction, Machine Learning, Electronic Health Records, Word Embeddings, Clinical Concept Similarity, Text Mining

## Abstract

**Background:** Electronic health records (EHRs) consist of both structured data (e.g., diagnostic codes) and unstructured data (e.g., clinical notes). It’s commonly believed that unstructured clinical narratives provide more comprehensive information. However, this assumption often lacks large-scale validation and direct validation methods.

**Objective:** This study aims to quantitatively compare the information in structured and unstructured EHR data and directly validate whether unstructured data offers more extensive information across a patient population.

**Methods:** We analyzed both structured and unstructured data from patient records and visits in a large Dutch primary care EHR database between January 2021 and January 2024. Clinical concepts were identified from free-text notes using an extraction framework tailored for Dutch and compared with concepts from structured data. Concept embeddings were generated to measure semantic similarity between structured and extracted concepts through cosine similarity. A similarity threshold was systematically determined via annotated matches and minimized weighted Gini impurity. We then quantified the concept overlap between structured and unstructured data across various concept domains and patient populations.

**Results:** In a population of 1.8 million patients, 42% of structured concepts in patient records and 25% in individual visits had similar matches in unstructured data. Conversely, only 13% of extracted concepts from records and 7% from visits had similar structured counterparts. Condition concepts had the highest overlap, followed by measurements and drug concepts. Subpopulation visits, such as those with chronic conditions or psychological disorders, showed different proportions of data overlap, indicating varied reliance on structured versus unstructured data across clinical contexts.

**Conclusions:** Our study demonstrates the feasibility of quantifying the information difference between structured and unstructured data, showing that the unstructured data provides important additional information in the studied database and populations. Despite some limitations, our proposed methodology proves versatile, and its application can lead to more robust and insightful observational clinical research.

## INTRODUCTION

Electronic Health Records (EHRs), originally designed for clinical documentation and administration, are now increasingly used in observational research [1,2], supporting various types of studies, including case studies, patient cohort characterizations, and clinical prediction modeling. EHR data is generally recorded in two forms: structured and unstructured data. Structured data includes clinical codes for documenting clinical events, such as diagnoses, medications, procedures, and measurements. Structured data is particularly suitable for observational research due to its consistent meaning, tabular format, and standardized vocabulary of codes. Unstructured data consists of free-text clinical notes, which can provide detailed descriptions capturing the nuances of patient care, such as physician observations, patient histories, diagnostic impressions, and discharge summaries. Although rich in contextual information, unstructured data poses challenges for direct analysis because of its variability and lack of standardization [3,4]. Consequently, extracting meaningful information from unstructured data requires significant investment in manual labor, computational resources, and time.

It is commonly assumed that the text data contains more detailed and extensive information than structured data, based on the often-reported claim – grounded in business-related data [5] – that 80% of EHR data is unstructured [3,4,6-8]. While this assumption may hold, its validity is influenced by factors like documentation quality and clinical context. For example, intensive care data is characterized by a high frequency of measurements, while psychiatric care relies more on textual narratives. Many studies explored the added value of information from text by comparing analyses with and without it [9-15], indirectly validating this assumption. However, even if the assumption holds, it initially remains uncertain to what extent the information in the text data matches or complements the structured data.

Understanding the quantity and differences in the information available in structured and unstructured data for a specific database offers several advantages for observational clinical research. First, it aids study design by identifying the most abundant and reliable data types, enabling researchers to formulate feasible hypotheses and research questions. Second, it allows for more effective allocation of human and computational resources by focusing efforts where they are most needed. Third, knowing the balance between structured and unstructured data helps researchers prioritize according to the study’s specific needs. Finally, it highlights gaps or unique aspects of the data, facilitating the exploration of underutilized research opportunities.

Comparing the information from structured and unstructured data involves various measures, such as quantity and content. While structured data points can be counted and unstructured data quantified by individual words or extracted concepts, comparing content similarity is more challenging. The core meaning of both structured codes and unstructured text lies in their semantic content. Evaluating the information distance between two concepts or texts requires comparing their semantic meanings, a task commonly addressed in natural language processing through semantic similarity measures. Modern approaches often utilize word embedding models to generate concept embeddings, which are used in applications like biomedical ontology matching [16,17] and concept normalization [18-20]. Specialized embedding models, such as SapBERT [21] and BioLORD [22], have been developed for this purpose and provide the opportunity to measure the information difference between structured and unstructured data.

To our knowledge, directly assessing the information difference between structured and unstructured data in a database has not been studied. Our study aims to quantitatively compare the information coded by general practitioners (GPs) with the information documented in free-text notes, using data from a large Dutch GP database. We extracted clinical concepts from unstructured text and used concept embeddings to calculate their similarity with the structured concepts. After determining a similarity threshold, we estimated the difference and overlap of information between structured and unstructured data.

## MATERIALS & METHODS

### Database and setting

This study utilized the Integrated Primary Care Information (IPCI) database [23], a longitudinal EHR database of Dutch GPs. IPCI contains records of 2.9 million patients with a median follow-up of 4.8 years, spanning from 1993 to 2024. The database is standardized using the Observational Medical Outcomes Partnership Common Data Model (OMOP CDM) [24]. Eligible participants in our study dataset included all patients recorded in the database from January 2021 to January 2024. The study received approval from the IPCI governance board under code 2023–04.

### Methodological setup

The methods consist of four main parts, visualized in Figure 1, each described in detail in the following sections. First, we extracted both structured and unstructured data for each eligible patient and applied a concept extraction framework to extract clinical concepts from the free-text notes. Second, we applied two different data grouping methods to the population. Third, we used pre-trained multilingual concept embeddings to calculate the similarities between the structured and extracted concepts. Finally, we annotated a sample of concept similarity matches to determine a similarity score threshold and quantified the data similarity in the database.

**Figure 1.**
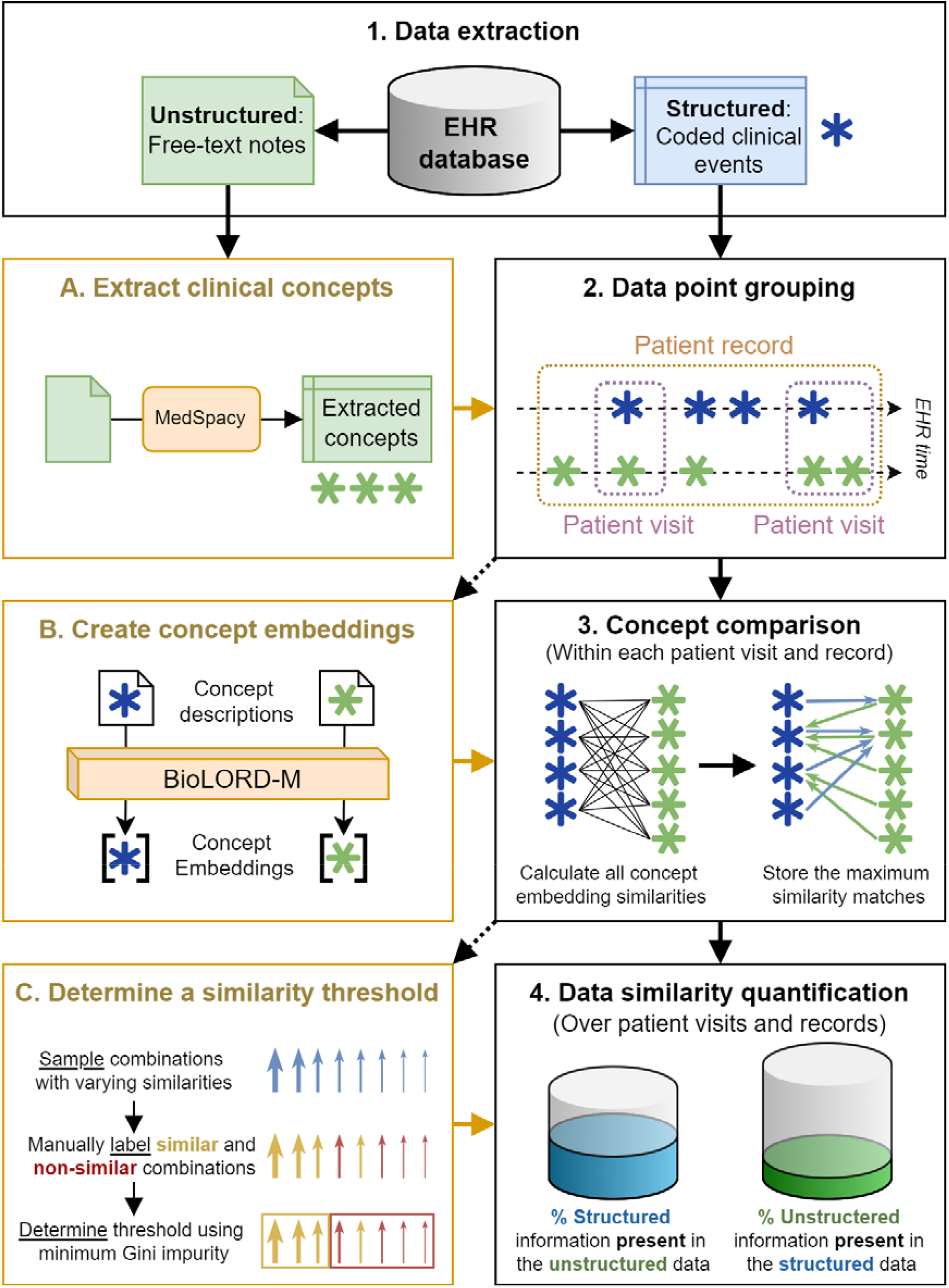
Visualization of the methodological setup. Steps 1 to 4 outline the primary process, including data extraction, data point grouping, concept comparison, and data similarity quantification. Steps A, B, and C detail additional processes, specifically the extraction of clinical concepts, the creation of concept embeddings, and the determination of a similarity threshold.

### Data extraction

We extracted all structured and unstructured data points for each eligible patient (Step 1 in Figure 1). Structured data includes conditions, procedures, prescriptions, measurements, and observations. The coding systems used, such as the International Classification of Primary Care (ICPC-1), are listed in Supplementary Table 1. Unstructured data consists of three types of notes: subjective, objective, assessment, and plan (SOAP) notes from consultations; referral and communication notes with secondary care; and other notes from free-text fields in the EHR system, primarily accompanying condition codes.

### Clinical concept extraction

Structured coded clinical events are considered single data points, while free-text notes can contain multiple pieces of information embedded in the narrative. To compare these, we extracted individual data points from unstructured text using clinical named entity recognition and linking, generally known as clinical concept extraction (Step A in Figure 1). We used MedSpacy [25], a toolkit that extracts clinical concepts based on a reference thesaurus such as the Unified Medical Language System (UMLS) and detects contextual modifiers using language-specific rules. We used a version of MedSpacy adapted for Dutch [10,26], incorporating all Dutch vocabularies from UMLS and replacing the English Systematized Nomenclature of Medicine Clinical Terms (SNOMED CT) with the Dutch translation [27]. We employed Dutch context rules to detect qualifiers such as negation, temporality, and experiencer. The extracted UMLS concepts were mapped to concept domains such as observation, condition, measurement, drug, and procedure in the OMOP standardized vocabulary, which contains the majority of UMLS vocabularies, for easy comparison with the structured data.

### Data point grouping

Given the longitudinal nature of GP EHR data, we used two grouping methods for comparison: by patient visit and by patient record (Step 2 in Figure 1). Grouping by visit considers data points recorded simultaneously during a patient visit, providing a natural basis for comparison, while grouping by record includes additional information recorded outside visits, such as lab results and secondary care communications, allowing for a broader comparison.

### Concept comparison

Each structured or extracted data point is represented by a single clinical concept. We compared the cartesian product of structured and extracted concepts within each group (Step 3 in Figure 1). For example, a visit with 1 coded condition and 1 prescription, along with a SOAP note containing 5 extracted concepts, results in 10 comparisons. While we could count exact concept matches, two issues arise. First, the compared concepts can be very similar but not identical, for example, the ICPC-1 code “Fracture: hand/foot bone” (L74) and the SNOMED CT concept “Closed fracture of hand” (704005005). Second, the concept vocabularies differ, as the GPs choose from a limited set of codes, while concept extraction uses the complete UMLS.

To overcome these issues, we used concept embeddings for fuzzy matching, enabling us to measure semantic similarities and recognize similar concepts. Cosine similarity, which measures the angle between two embedding vectors, was employed to calculate similarities, ranging from -1 (strongly opposite) to 1 (very similar). For each structured and extracted concept, we stored only the highest similarity match, as we are only interested in finding the most similar concept in the other data type.

### Concept embeddings

We used the BioLORD-2023-M pre-trained sentence transformer model [22] to create the concept embeddings, as visualized in step B of Figure 1. By inputting the concept description, from either UMLS for the extracted concepts or form the source vocabulary for the structured concepts, the model generates a dense 768-dimensional vector, allowing us to create embeddings for both structured and extracted concepts. For concepts with multiple descriptions or synonyms, we calculated an embedding for each and averaged them to create one comprehensive concept representation. For structured observations and measurements, we included the unit and value in the description to enrich the embedding. The model’s multilingual capability enabled us to embed concept descriptions in both Dutch and English within the same latent space.

### Similarity threshold determination

To quantify the information difference between structured and extracted concepts, we needed to define a threshold for concept similarity. Since concept similarity depends on the nature of the embeddings, we developed a systematic method to determine this threshold, as visualized in step C of Figure 1. First, we randomly selected concept pairs at various similarity levels, ranging from a similarity score of 0.35 to 1, with samples taken at 0.05 intervals. This sampling was done for both structured and extracted concepts across patient visits and records, ensuring each concept domain was represented by sampling five concepts per domain. Next, we manually annotated the concept pairs as either similar or non-similar. Using these annotations, we determined the threshold at which the weighted Gini impurity [28] of the split between similar and non-similar matches was lowest.

### Data similarity quantification

Using a similarity threshold and the most similar counterpart for each structured and extracted concept, we determined the number of structured concepts found in free-text (structured-to-unstructured) and the number of extracted concepts that were coded in the structured data (unstructured-to-structured), as shown in Step 4 of Figure 1. It is important to note that these counts are not reciprocal since we consider the maximum similarity per concept. For example, multiple extracted concepts from a patient visit text may be highly similar to a single structured concept, but we only compare the structured concept to its most similar extracted counterpart to determine its presence in the text, not the frequency. We calculated these counts and their percentages across the entire set of concepts, as well as within different concept domains, to explore domain-specific differences. For extracted concepts, we only included those without context modifiers.

### Subpopulation comparison

While observing data similarity across the entire population or all GP visits is insightful, applying this method to smaller subpopulations may provide further detail. We defined three subpopulations based on different types of clinical events: visits for chronic disorder (type 2 diabetes mellitus), acute event (COVID-19 vaccination), and psychological disorder (depression), as detailed in Supplementary Table 2. We then quantified the similarity between structured and unstructured data for these subpopulations, similar to the full population.

## RESULTS

### Population and data characteristics

Table 1 presents the population and data characteristics for patient records and visits, including details per observation. Additionally, characteristics are listed for three subpopulations. Key differences between patient visits and records are evident. First, each patient has one complete record but can have multiple visits. Second, the total number of structured events and their median values indicate that many events are recorded outside GP visits, as their total numbers are roughly three times higher in records. Similarly, the total number of clinical notes is twice as high in records compared to visits alone. Third, more extracted concepts per structured event are generally found in patient visits, but the median number of concepts per note is the same.

**Table 1.** Population and data characteristics for patient records, patient visits, and three subpopulation-specific visits.

|  | Patient records | Patient visits | Covid-19<br>vaccination visits | Depression visits | Diabetes visits |
| --- | --- | --- | --- | --- | --- |
| **Total numbers** |  |  |  |  |  |
| No. patients | 1,794,209 | 1,507,473 | 62,724 | 20,519 | 51,032 |
| No. observations | 1,794,209 | 13,995,524 | 84,484 | 63,957 | 324,698 |
| Ratio no. observations / no. patients | 1 | 9.28 | 1.35 | 3.12 | 6.36 |
| No. structured events | 132,287,566 | 41,232,407 | 281,606 | 118,351 | 4,513,756 |
| No. free-text notes | 95,790,377 | 47,915,131 | 188,318 | 216,449 | 1,253,418 |
| No. extracted concepts | 635,232,943 | 245,052,660 | 688,420 | 1,374,430 | 5,329,978 |
| Ratio extracted concepts / structured events | 4.80 | 5.94 | 2.44 | 11.61 | 1.18 |
| **Statistics per observation** |  |  |  |  |  |
| Percentage male | 49% | 49% | 45% | 33% | 51% |
| Median age | 39 | 39 | 62 | 49 | 68 |
| Median no. structured events | 33 | 2 | 2 | 1 | 6 |
| Median no. free-text notes | 35 | 3 | 2 | 3 | 3 |
| Median no. extracted concepts | 172 | 12 | 5 | 15 | 12 |
| Median no. concepts per note | 3 | 3 | 2 | 3 | 3 |

Characteristics for visits across different clinical events also show clear contrasts. As expected, visits regarding diabetes presented the most visits per patient, while for Covid-19 vaccination the number of visits per patient was the least. Furthermore, diabetes visits contain much structured and unstructured data, whereas depression visits rely mostly on unstructured data with a large median number of extracted concepts per visit and few structured events. Demographic differences are also notable: diabetes and vaccination populations are similar, but the depression population consists of younger females.

### Determining the similarity threshold

We annotated 1,764 matches in four concept samples: structured-to-unstructured and unstructured-to-structured in both patient record and visit populations. For transparency and reproducibility, the annotated samples are available in Supplementary Table 3. Figure 2B visualizes some example extracted concepts matched with increasing similarity to the structured concepts Sars-CoV-2 (COVID-19) and Hypertension. The weighted Gini impurity calculated over the binary annotated matches at different similarity thresholds is presented in Figure 2A. We found the minimum impurity of each of the four samples between thresholds of 0.55 and 0.65. To be conservative, we set the threshold at 0.65 for data similarity quantification.

**Figure 2.**
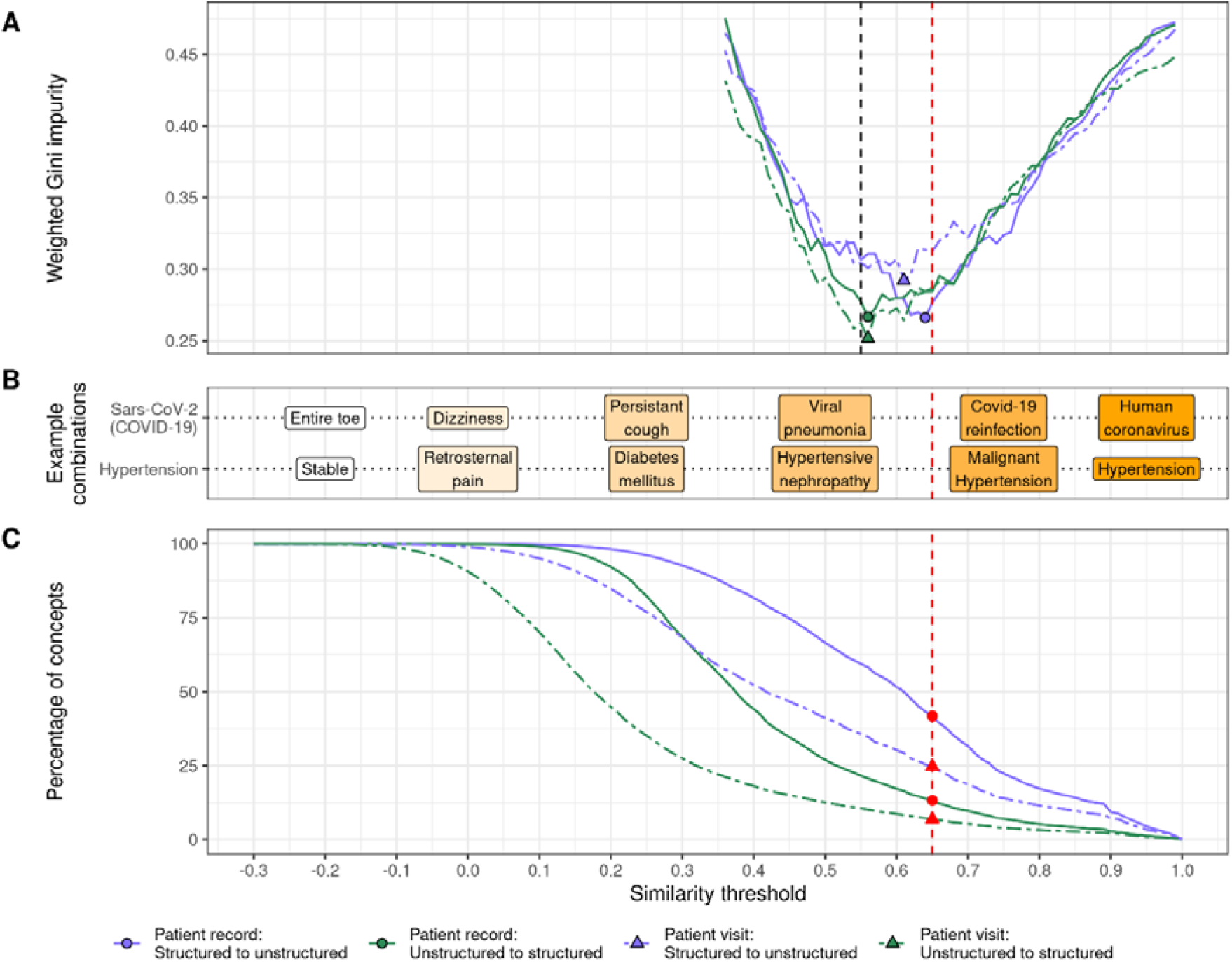
Analysis across similarity thresholds: A. The weighted Gini impurity measures the level of impurity in the annotated concept combinations in the four samples over the different thresholds. The points indicate the minimum impurity in each sample, and the dashed lines represent their lower bound (0.55) and upper bound (0.65). B. Examples of extracted concepts matched to the structured concepts of Sars-CoV-2 (COVID-19) and Hypertension across different similarity thresholds. C. The percentage of structured concepts matched the extracted concepts (blue) over the range of similarity thresholds and vice versa (green) for both patient records and visits. The red dashed line in all figures depicts the determined similarity threshold value of 0.65.

### Similarity of structured and unstructured data

The percentage of structured concepts that have a similar match to an extracted concept and vice versa over various similarity thresholds is visualized in Figure 2C for both patient visits and records. At the similarity threshold of 0.65, indicated by the red line in Figure 2, 42% of structured concepts in patient records are similar to a concept extracted from text, compared to 25% for visits. In contrast, only 13% of extracted concepts are similar to a structured concept in patient records, and 7% in visits (Figure 3A). This indicates that information in the structured data is more likely to be present in the unstructured data than vice versa. The difference between patient records and visits can be explained by the number of available concepts for matching the entire record versus a single visit.

**Figure 3.**
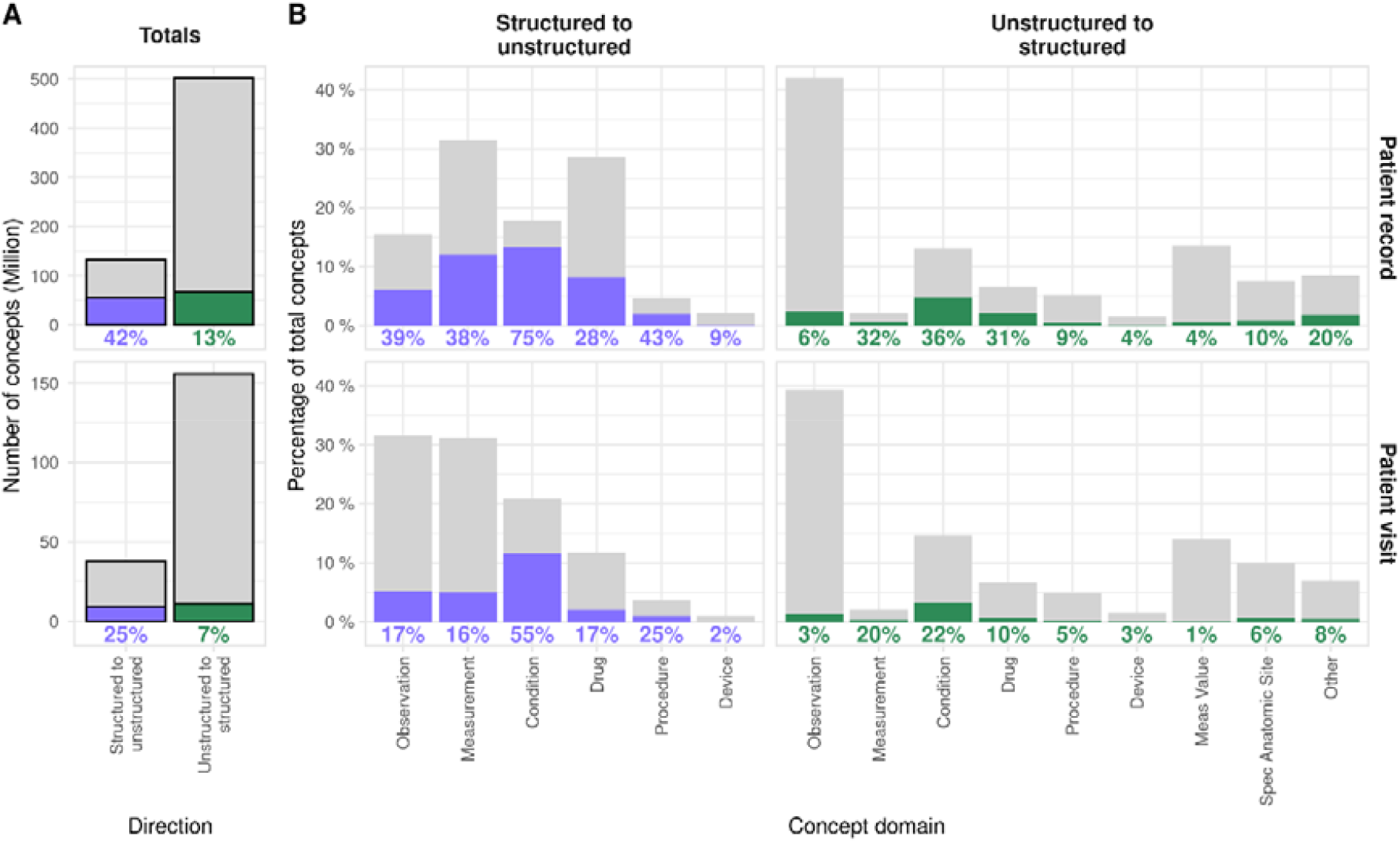
Comparison of structured and unstructured concepts in patient records and visits. (A) Total number of structured and unstructured concepts (grey), along with the number of concepts matched at a similarity threshold of 0.65 (blue for structured to unstructured, green for unstructured to structured). The percentage of matched concepts is listed under the chart. (B) The proportion of each concept domain contributed to the total number of structured and unstructured concepts (grey), along with the proportion of concepts in each domain matched to the other data type.

Figure 3B presents the counts and overlap percentages for different domains of structured and extracted concepts. Primarily structured conditions are often also in the text, with 75% for patient records and 55% for visits. Similarly, extracted condition concepts are also most often matched with structured concepts, with 36% in patient records and 22% in visits. Other concept domains, such as measurements and drugs, show a relatively high overlap in both structured and unstructured data as well.

Interestingly, differences in concept domain numbers between patient visits and records are larger for structured concepts than extracted concepts. For example, the proportion of structured observation concepts is 31% in patient visits but 15% in patient records, and the reverse is true for drug concepts. However, this difference is not observed in extracted concepts.

### Differences between subpopulations

Figure 4A presents the total concept overlap at the selected threshold for the three subpopulations, showing ratios of structured and extracted concepts as seen in Table 1. While the proportion of structured data overlapping with unstructured data and vice versa is similar for the vaccination and diabetes populations, the depression population shows a much higher proportion of structured data matched to concepts in the text (40%) and a lower proportion of extracted concepts matched to structured concepts (5%). Figure 4B shows different proportions of concept domains in the structured data across the three populations. However, in the extracted concepts, the proportions are relatively similar. The data similarity results are consistent with the total population, with conditions, drugs, and measurement concepts showing the highest overlap between structured and unstructured data.

**Figure 4.**
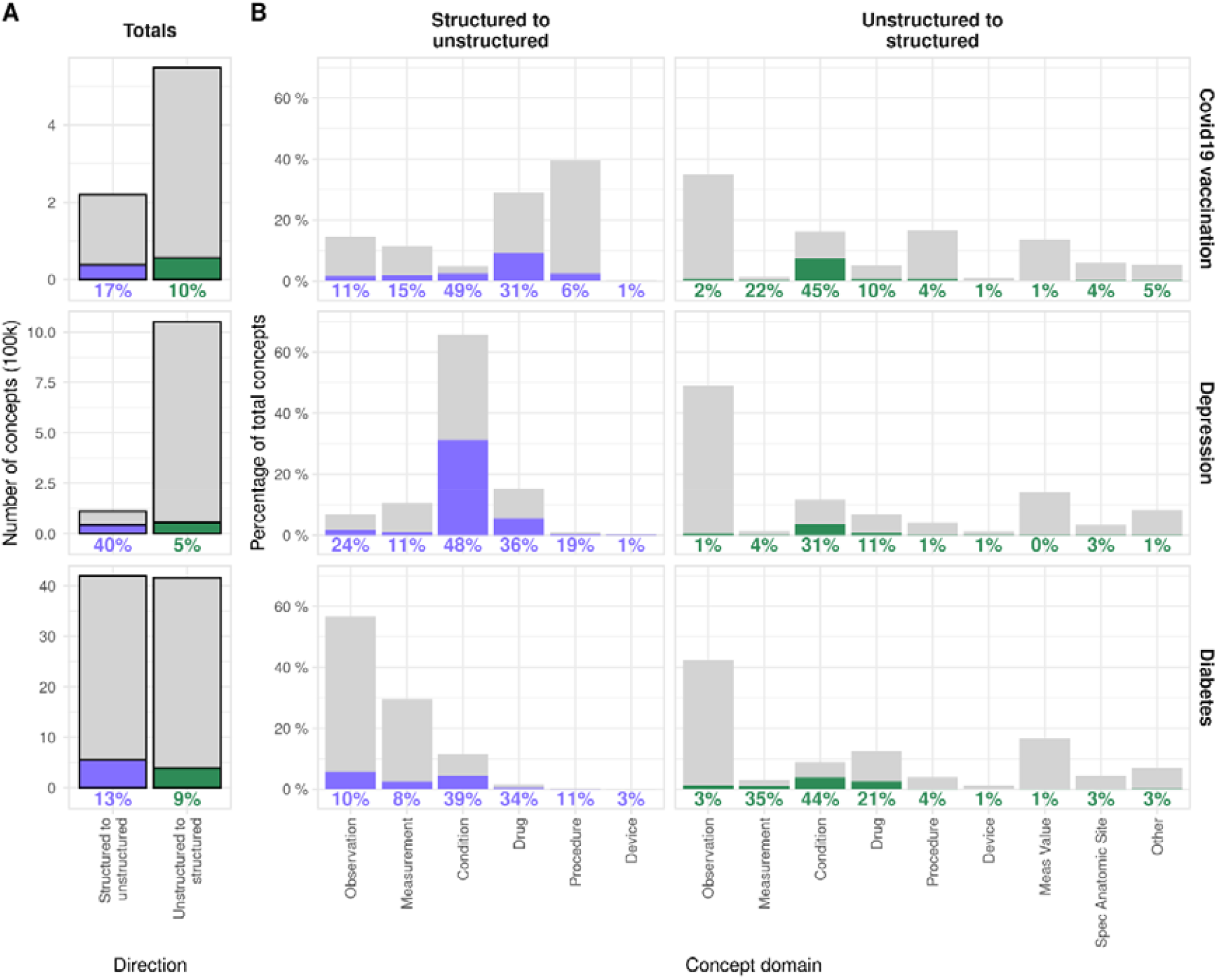
Comparison of structured and unstructured concepts in the three subpopulation visits. (A) Total number of structured and unstructured concepts (grey), along with the number of concepts matched at a similarity threshold of 0.65 (blue for structured to unstructured, green for unstructured to structured). The percentage of matched concepts is listed under the chart. (B) The proportion of each concept domain contributed to the total number of structured and unstructured concepts (grey), along with the proportion of concepts in each domain matched to the other data type.

## DISCUSSION

### Comparing structured and unstructured clinical data

This study explored the feasibility of quantifying the information difference between structured and unstructured data in a large primary care EHR database. We utilized concept embeddings to measure the similarity between structured clinical event concepts and concepts extracted from free-text narratives. By systematically determining a similarity threshold, we found that a substantial proportion of structured information is also present in unstructured data, while only a small portion of unstructured information is reflected in structured data. This indicates that most concepts in one data type do not match those in the other, suggesting that the information in structured and unstructured data is highly complementary.

Furthermore, we observed that condition concepts had the largest overlap between structured and unstructured data, followed by measurements and drug concepts. These results were consistent across different data point grouping methods, by patient record or visit. We also quantified the information difference in smaller subpopulations of patients with specific diseases or procedures. Differences in overlap proportions were evident between these subpopulations, but the concept domains with the largest overlap were the same as in the full population. Overall, our findings validate the often-assumed notion that unstructured data contains more extensive information than structured data in this specific database. Most importantly, we prove the feasibility of quantifying the information difference in an EHR database using a combination of clinical concept extraction methods [25], high-dimensional clinical concept embeddings for similarity measurement [22], and annotating a small number of matched concepts to determine the appropriate similarity threshold.

### Strengths and limitations

There are several limitations to our methodology. The results depend heavily on three factors: the type and quality of concept extraction, the type of concept embeddings used for determining concept similarity, and the chosen similarity threshold. Firstly, the performance of named entity recognition and entity linking is crucial for extracting all relevant information from unstructured data. Imperfect extraction can lead to missed concepts (false negatives) that cannot be matched to structured concepts and wrongly extracted concepts (false positives) that distort the results. The MedSpacy extraction framework used in this work was validated for different languages and demonstrates good performance [25,26]. It was chosen for its availability in Dutch, its versatility with ontologies, and its extraction speed. Secondly, different types of word and concept embeddings may vary in their ability to distinguish between similar and non-similar clinical concepts, affecting the distribution of similarity scores. While other, more complex embeddings might improve similarity discrimination, we only tested the multilingual BioLord-2023-M embedding model [22], as it was specifically trained to produce meaningful representations for clinical sentences and biomedical concepts, making it perfectly suited to our application. Thirdly, the chosen similarity threshold impacts the results. Setting this threshold can be subjective and heavily depends on the concept embeddings. Therefore, we used a systematic approach using the minimization of the Gini impurity to determine the threshold and we published the annotated concept matches for transparency. Lastly, data grouping poses a challenge. We used two grouping methods to explore differences, but more sophisticated approaches might be necessary depending on the research question. For instance, data recorded outside patient visits may be assigned to the nearest visit.

Overall, the strength of this study lies in enhancing our understanding of the information overlap between structured and unstructured clinical data within a large GP EHR database covering 1.8 million patients. Our methodology of using concept embeddings to calculate the similarity between clinical concepts offers a versatile and language-independent solution, demonstrated here for Dutch, ensuring accurate comparisons by capturing nuances in clinical terminology.

### Future work

Future research in quantifying the data similarity or difference between different data types could explore more advanced concept extraction and embedding techniques or alternative similarity measurements beyond cosine similarity. Our study focused on individual concepts for comparison, but combining multiple concepts might yield higher similarity matches. Investigating the use of concept n-grams and further incorporating context modifiers of the extracted concepts could be beneficial. We applied concept extraction before embedding to retain the granular meaning of individual events within a document. This approach ensures that specific events—such as symptoms, prescriptions, and procedures—are not lost in broader document or sentence embeddings. Future research could explore directly comparing structured concepts and free text without prior clinical concept extraction to enhance the method’s reliability and applicability.

We used a single similarity threshold across all concept domains and populations. Future work could involve establishing separate thresholds for each concept domain and population for a more refined comparison. The potential applications of our methodology extend beyond the clinical domain. With the appropriate information extraction framework and embedding model, our approach could be adapted to various settings.

## CONCLUSION

Our study aimed to assess the feasibility of quantifying the information difference between structured and unstructured data in EHR databases. Within a large Dutch primary care EHR database, we successfully demonstrated that unstructured data provides more extensive information than structured data, quantitatively validating this prevailing assumption. By leveraging concept embeddings to measure semantic similarity between structured concepts and those extracted from free-text narratives, we found that a significant portion of structured information is present in unstructured data, while the reverse occurs much less frequently. Notably, concept domains such as conditions, measurements, and drugs exhibited the largest overlap. Despite limitations related to the performance of concept extraction, the type of embeddings used, and the determination of similarity thresholds, our methodology is versatile and was applied across different data grouping methods and subpopulations. The exploration of more sophisticated techniques could further enhance the accuracy and applicability of this approach. Understanding the extent and nature of information in structured and unstructured data within a database can enhance study design, research exploration, resource allocation, and data prioritization, ultimately leading to more robust and insightful observational clinical research.

## Data Availability

The annotated concept matches are available as supplementary material.

## AUTHOR STATEMENT

T.S. proposed the methodology, designed, and implemented the study protocol, and performed the data analysis. J.K., E.M., and P.R. provided critical feedback, helped interpret the results, and shaped the research and analysis. T.S. wrote the article with valuable input from all other authors.

## ACKNOWLEDGMENTS

There is nothing to declare.

## REFERENCES

1. Knevel R, Liao KP. From real-world electronic health record data to real-world results using artificial intelligence. Annals of the Rheumatic Diseases 2023;82(3):306–11.

2. Reps JM, Schuemie MJ, Suchard MA, et al. Design and implementation of a standardized framework to generate and evaluate patient-level prediction models using observational healthcare data. Journal of the American Medical Informatics Association 2018;25(8):969–75.

3. Assale M, Dui LG, Cina A, et al. The revival of the notes field: leveraging the unstructured content in electronic health records. Frontiers in medicine 2019;6:66.

4. Li I, Pan J, Goldwasser J, et al. Neural natural language processing for unstructured data in electronic health records: a review. Computer Science Review 2022;46:100511.

5. Murdoch TB, Detsky AS. The inevitable application of big data to health care. Jama 2013;309(13):1351–52.

6. Chiu C-C, Wu C-M, Chien T-N, et al. Integrating structured and unstructured EHR data for predicting mortality by machine learning and latent Dirichlet allocation method. International Journal of Environmental Research and Public Health 2023;20(5):4340.

7. Hashir M, Sawhney R. Towards unstructured mortality prediction with free-text clinical notes. Journal of biomedical informatics 2020;108:103489.

8. Kong H-J. Managing unstructured big data in healthcare system. Healthcare informatics research 2019;25(1):1–2.

9. Seinen TM, Fridgeirsson EA, Ioannou S, et al. Use of unstructured text in prognostic clinical prediction models: a systematic review. Journal of the American Medical Informatics Association 2022;29(7):1292–302.

10. Seinen TM, Kors JA, van Mulligen EM, et al. The added value of text from Dutch general practitioner notes in predictive modeling. Journal of the American Medical Informatics Association 2023;30(12):1973–84.

11. Kharrazi H, Anzaldi LJ, Hernandez L, et al. The value of unstructured electronic health record data in geriatric syndrome case identification. Journal of the American Geriatrics Society 2018;66(8):1499–507.

12. Zhang D, Yin C, Zeng J, et al. Combining structured and unstructured data for predictive models: a deep learning approach. BMC medical informatics and decision making 2020;20:1–11.

13. Gan S, Kim C, Chang J, et al. Enhancing readmission prediction models by integrating insights from home healthcare notes: Retrospective cohort study. International Journal of Nursing Studies 2024;158:104850.

14. Marafino BJ, Park M, Davies JM, et al. Validation of prediction models for critical care outcomes using natural language processing of electronic health record data. JAMA network open 2018;1(8):e185097–e97.

15. Tsui FR, Shi L, Ruiz V, et al. Natural language processing and machine learning of electronic health records for prediction of first-time suicide attempts. JAMIA open 2021;4(1):ooab011.

16. Park J, Kim K, Hwang W, et al. Concept embedding to measure semantic relatedness for biomedical information ontologies. Journal of biomedical informatics 2019;94:103182.

17. Zhang Y, Wang X, Lai S, et al. Ontology matching with word embeddings. Chinese Computational Linguistics and Natural Language Processing Based on Naturally Annotated Big Data: 13th China National Conference, CCL 2014, and Second International Symposium, NLP-NABD 2014, Wuhan, China, October 18-19, 2014. Proceedings; 2014. Springer.

18. Abdulnazar A, Kreuzthaler M, Roller R, et al. SapBERT-based medical concept normalization using SNOMED CT. Caring is Sharing–Exploiting the Value in Data for Health and Innovation: IOS Press, 2023:825–26.

19. Zahra FA, Kate RJ. Obtaining clinical term embeddings from SNOMED CT ontology. Journal of Biomedical Informatics 2024;149:104560.

20. Xu D, Miller T. A simple neural vector space model for medical concept normalization using concept embeddings. Journal of biomedical informatics 2022;130:104080.

21. Liu F, Vulić I, Korhonen A, et al. Learning domain-specialised representations for cross-lingual biomedical entity linking. arXiv preprint arXiv:2105.14398 2021.

22. Remy F, Demuynck K, Demeester T. BioLORD-2023: semantic textual representations fusing large language models and clinical knowledge graph insights. Journal of the American Medical Informatics Association 2024:ocae029.

23. de Ridder MA, de Wilde M, de Ben C, et al. Data resource profile: the integrated primary care information (IPCI) database, The Netherlands. International Journal of Epidemiology 2022;51(6):e314–e23.

24. Hripcsak G, Duke JD, Shah NH, et al. Observational Health Data Sciences and Informatics (OHDSI): opportunities for observational researchers. MEDINFO 2015: eHealth-enabled Health: IOS Press, 2015:574–78.

25. Eyre H, Chapman AB, Peterson KS, et al. Launching into clinical space with medspaCy: a new clinical text processing toolkit in Python. AMIA Annual Symposium Proceedings; 2021. American Medical Informatics Association.

26. Seinen TM, Kors JA, van Mulligen EM, et al. Annotation-preserving machine translation of English corpora to validate Dutch clinical concept extraction tools. medRxiv 2024:2024.03. 14.24304289.

27. SNOMED National Release Centre of the Netherlands. 2024. https://www.snomed.org/member/netherlands (accessed September 25, 2024).

28. Nembrini S, König IR, Wright MN. The revival of the Gini importance? Bioinformatics 2018;34(21):3711–18.

