## Supplementary for "Structured Codes and Free-Text Notes: Measuring Information Complementarity in Electronic Health Records"

**SUPPLEMENTARY MATERIAL**

**Supplementary tables**

*Supplementary Table 1. Concept vocabularies used for the structured data in the IPCI database*

| **Structured data domain** | **Vocabulary coding system** |
| --- | --- |
| Conditions | ICPC-1 |
| Procedures | ICPC-1 |
|  | Vektis |
| Observations | NHG-45 |
|  | ICPC-1 |
|  | Vektis |
| Measurements | NHG-45 |
|  | ICPC-1 |
| Medication | Z-index |
| Device | Z-index |
|  | ICPC-1 |

ICPC-1 – International Classification of Primary Care 1

NHG-45 – Dutch College of General Practitioners, Table 45: Diagnostic determinations

Vektis – Insurance codes from the executive organization of Dutch health insurers.

Z-index – Vocabulary with detailed information on all medicines and devices available in the Netherlands.

*Supplementary Table 2. Definitions of the three subpopulations*

| **Subpopulation** | **Definition** | **ICPC-1 code** | **SNOMED CT concept code** | **OMOP standardized vocabulary concept id** |
| --- | --- | --- | --- | --- |
| Type 2 diabetes mellitus | A visit with condition occurrence of *Type 2 diabetes mellitus* | T90.02 | 44054006 | 201826 |
| COVID-19 vaccination | A visit with procedure occurrence of *COVID-19 vaccination* | R44.91 | 127785005 | 4132855 |
| Depression | A visit with condition occurrence of *Depressive disorder* | P76 | 35489007 | 440383 |

*Supplementary Table 3. Table with the annotated concept matches.*

CSV file: SupplementaryTable3_annotated_concept_matches.csv
